# Gene expression based molecular test proves clinical validity as diagnostic aid for the differential diagnosis of psoriasis and eczema in formalin fixed and paraffin embedded tissue

**DOI:** 10.1101/2022.05.18.22275097

**Authors:** Felix Fischer, Sophie Roenneberg, Larissa Graner, Franziska Schlenker, Roland Zengerle, Fabian J. Theis, Carsten B. Schmidt-Weber, Tilo Biedermann, Felix Lauffer, Natalie Garzorz-Stark, Stefanie Eyerich

## Abstract

Highly specific and efficient drugs have been developed during the last two decades to treat non-communicable chronic inflammatory skin diseases (ncISD). Due to their specificity, these drugs are asking for precise diagnostic measures to attribute the most efficient treatment to each patient. Diagnosis, however, is complicated by the complex pathogenesis of ncISD and their clinical and histological overlap. Especially, precise diagnosis of psoriasis and eczema is difficult in special cases and molecular diagnostic tools need to be developed to support gold standard diagnosis of patients. In this line, we have developed a real-time based molecular classifier to distinguish psoriasis from eczema in RNA-later fixed skin samples. However, this type of skin sample is not regularly used in routine diagnostics. Therefore, we evaluated if the combination of *NOS2* and *CCL27* expression in lesional skin can be transferred to formalin-fixed paraffin embedded (FFPE) tissue. We present a FFPE-based molecular classifier (MC) that determines the probability for psoriasis with a specificity and sensitivity of 100% and 92%, respectively, and an area under the curve (AUC) of 0.97 delivering comparable results to the RNA-later based MC. The probability for psoriasis as well as the PCR result of *NOS2* expression correlated positive with disease hallmarks of psoriasis and negative with eczema hallmarks. This tool now offers broad usage in pathology laboratories and can support diagnostic decision making on a molecular level.

## Introduction

Psoriasis and eczema are one of the most prevalent and relevant skin diseases worldwide with an incidence rate for both diseases together of at least 2,000/100,000 (Bylund, von Kobyletzki, Svalstedt, & Svensson, 2020; Thyssen, Johansen, Linneberg, & Menne, 2010; Vena et al., 2010). Although more than a dozen of specific immune-modulating therapies including biologicals and small molecules have been made available to treat patients more efficiently than ever before over the last decade (Tizek et al., 2021), the ultimate breakthrough has yet to arise due to missing accurate diagnostics tools. Diagnosis of both diseases largely depends on a subjective visual exam and histopathological examination. Due to phenotypical overlaps of both diseases up to 50 % of cases are misdiagnosed (Kolesnik, Franke, Lux, Quist, & Gollnick, 2018). To close this important diagnostic gap, a gene expression based classifier using *NOS2* and *CCL27* as marker genes has been proposed and its clinical validity has been examined and proven in various patient cohorts (Bentz et al., 2021; Brunner et al., 2018; Garzorz-Stark et al., 2016; Quaranta et al., 2014; Stoffel et al., 2018). Renert-Yuval et al. list the *NOS/CCL27* classifier as the only accepted disease classifier for psoriasis and eczema (Renert-Yuval et al., 2021). Here, we applied the molecular classifier in formalin-fixed paraffin embedded biopsy specimens of eczema and psoriasis to validate its clinical utility as diagnostic aid in pathology laboratories (n=107). We demonstrated that the FFPE-based classifier delivered the correct clinical and histopathological diagnosis on the level of RT-PCR with a test sensitivity and specificity of 92% and 100%, respectively. By opening FFPE samples to molecular diagnostics, this FFPE-based classifier can now be used in routine diagnostics and aid physician in their diagnostic decision making.

## Material and Methods

### Patients and material sampling

FFPE samples for this retrospective study were derived from the Biobank Biederstein and consisted of 107 patients (psoriasis, n=27; eczema, n=48, disease subtypes, n=18 and cases with contradictory diagnosis n=14)). Final diagnosis of patients was determined by at least two dermatologists taking a comprehensive view onto the clinical picture, family history, laboratory parameters and histological assessment. Characteristics of all patients are shown in Table 1. Patients treated with immune-efficient medication prior to material sampling were excluded from the study. The study was approved by the local ethical committee (project number 2773/10 and 515/17). Generated data and codes can be provided upon request.

**Table 1:** Patient characteristics

|  | n | age |  | n | age |
| --- | --- | --- | --- | --- | --- |
| *psoriasis )pso) (n=27)* |  |  | *aopic dermatitis (n=28)* |  |  |
| m | 20 | 49.7±12.56 | m | 13 | 47.69±25.82 |
| w | 7 | 65.86±10.12 | w | 15 | 55.67±29.08 |
| *psoriasis guttata (PG) (n=5)* |  |  | *nummular eczema (n=21)* |  |  |
| m | 2 | 36.5±5.5 | m | 10 | 62.6±15.32 |
| w | 3 | 40.23±3.86 | w | 11 | 72±14.25 |
| *generalized pustular psoriasis (GPP) (n=2)* |  |  | *dishidrotic eczema (DHE) (n=4)* |  |  |
| m | 1 | 55-60 | m | 1 | 70-75 |
| w | 1 | 25-30 | w | 3 | 55.67±19.29 |
| *psoriasis palmoplantaris (PP) (n=1)* |  |  | *hyperkeratotic-rhagadiiform eczema (HRE) (n=3)* |  |  |
| w | 1 | 40-45 | m | 1 | 60-65 |
| *psoriasis pustulosa palmoplantaris (PPP) (n=3)* |  |  | w | 2 | 70±21.21 |
| m | 2 | 42±9.9 | *unclear cases (n=14)* |  |  |
| w | 1 | 45-50 | m | 8 | 56.86±13.36 |
|  |  |  | w | 6 | 66±9.7 |

### Characteristics of FFPE tissue and isolation of total RNA

Skin biopsies (Ø3-6mm) from patients with a potential diagnosis of psoriasis or eczema were taken under local anesthesia by a dermatologist and immediately fixed in neutral buffered formalin for 8-48 hours at room temperature or 4°C. Fixed biopsies were paraffin embedded according to standard procedures (one biopsy per paraffin cassette). Unstained FFPE skin biopsies (<10 years of age) were used to isolate mRNA. Here, tissue sections with a thickness of 10 μm were generated from each FFPE skin sample. The first sections containing both epidermis and dermis were trimmed (at least 30μM) and discarded to avoid usage of degraded material. mRNA was extracted from in total three FFPE sections using a magnetic bead-based purification method according to the manufacturer’s protocol (InnuPREP MP Basic FFPE RNA Kit, IST Innuscreen, Germany). The mRNA eluates were analyzed immediately after the extraction by real-time PCR using specifc primer and hydrolysis probes.

### Primer design and Real-time PCR

For specific amplification of the genes of interests *NOS2* and *CCL27* as well as of the reference genes, *TBP* and *SDHAF2*, primers and hydrolysis probes were designed using the primer design tool PrimerBLAST from NCBI (https://www.ncbi.nlm.nih.gov/tools/primer-blast/). Oligonucleotides were synthesized by biomers (Germany). *P*robes were labeled with an Atto 647N fluorophore at the 5’-end and quenched with a BHQ2 quencher at the 3’-end.

Primer and probe sequences were as follows (forward, reverse and probe): 5’- TTCCTGGTTTGACTGTCCTTACC-3’, 5’- T7TTCACTGTGGGGCTTGC-3’ and 5’- CGGGGAGGCAGTGCAGCCAGC-3’ for *NOS2*; 5’- ACCCTACAGCAGCATTCCTAC-3’, 5’- AGCTTGTCTGAGAGTGGCTTTC-3’ and 5’- ACCCAGCACTGCCTGCTGTACTCAGCTC-3’ for *CCL27;* 5’- ATGGATCAGAACAACAGCCTGC-3’, 5’- CATTGGACTAAAGATAGGGATTCCG-3’ and 5’-TCAGGGCTTGGCCTCCCCTCAGGGTGCC-3’ for *TBP;* 5’- CGGTGTCTACAGTGTTCTCGAC-3’, 5’- GATGTCACACTGAGCAAAGGAG-3’ and 5’- CGCTGATGCTTGCTCTGTCAAGGCACAGCC-3’ for *SDHAF2*.

Real-time PCR reactions were carried out in 96-well plates using a Thermo Fisher Scientific one-step RT-PCR master mix. Thermocycling and fluorescence detection were performed in a qTower^3^ (Analytik Jena, Germany) and amplification results were analyzed using the qPCRsoft 4.1 software (Analytik Jena).

### FFPE-based molecular classifier (MC) built-up

The molecular classifier was built using L2 regularized logistic regression. The molecular classifier predicts the disease state (probability for psoriasis) directly from the normalized CT values of *NOS2* and *CCL27* normalized to the reference genes *SDHAF2* and *TBP*. For classification the psoriasis and eczema subtypes are aggregated to their respective high level labels (eczema or psoriasis). The model was calculated using LinearRegression implementation from scikit-learn with an L2 regularization strength of 100. In addition, the uncertainty of the model is estimated based on the predicted probability (cases with a predicted probability between 45% and 55% are predicted as undetermined). The model was trained on 77 patients with a clear diagnosis of psoriasis and eczema (atopic dermatitis (AD) and nummular eczema (NE)) and achieved an AUC of 0.97 +/- 0.06 on the training data. Furthermore, the model achieved a sensitivity = 91% and a specificity = 91% on the evaluation set which includes the training data plus 18 additional patients with prediagnosed subtypes of psoriasis and eczema.

### Correlation of FFPE-based MC probability and PCR results with clinical attributes

Pearson correlation coefficients were calculated to determine the correlation between the FFPE-based MC determined probabilities for psoriasis, the negative delta CT PCR results of *CCL27* and *NOS2* as well as histological parameters, morphology of the lesion, comorbidities, laboratory parameters and family history.

## Results

### The FFPE-based molecular classifier (MC) precisely separates eczema from psoriasis and efficiently identifies subtypes of both diseases

To evaluate the potential of FFPE samples for molecular diagnostics, 76 FFPE samples of lesional psoriasis (n=27) and eczema (atopic dermatitis n=28; nummular eczema n=21) were analyzed by probe-based real-time PCR for the expression of *NOS2* and *CCL27*. To calculate a FFPE-based molecular classifier (MC) for differentiation of psoriasis and eczema, a logistic regression model was fitted with a 5-times cross validation. This classifier was designed to predict the probability of psoriasis with a cut-off value of 55% (The range between 45% and 55% would no probability (undetermined). Test specificity for psoriasis was 100% with a sensitivity of 92% and an area under the ROC curve (AUC) of 0.97 +/- 0.06 (Fig. 1A). Two psoriasis patients were mis-classified, and two further psoriasis patients reached a probability between 45-55% and thereby could not be evaluated (Fig. 1B). In a next step, we used the FFPE-based MC to classify subtypes of psoriasis (psoriasis guttata n=5; psoriasis palmoplantaris n=1; generalized pustular psoriasis n=2; and psoriasis pustulosa palmoplantaris n=3) and eczema (dishidrotic eczema n=4; hyperkeratotic rhagadiforme eczema n=3) (Fig. 1B). Whereas all cases of psoriasis guttata, psoriasis palmoplantaris and psoriasis pustulosa pampoplantaris were diagnosed correctly by the FFPE-based MC, cases of dishidrotic and hyperkeratotic-rhagadiform eczema as well as generalized pustular psoriasis tended to be misdiagnosed.

**Figure 1:**
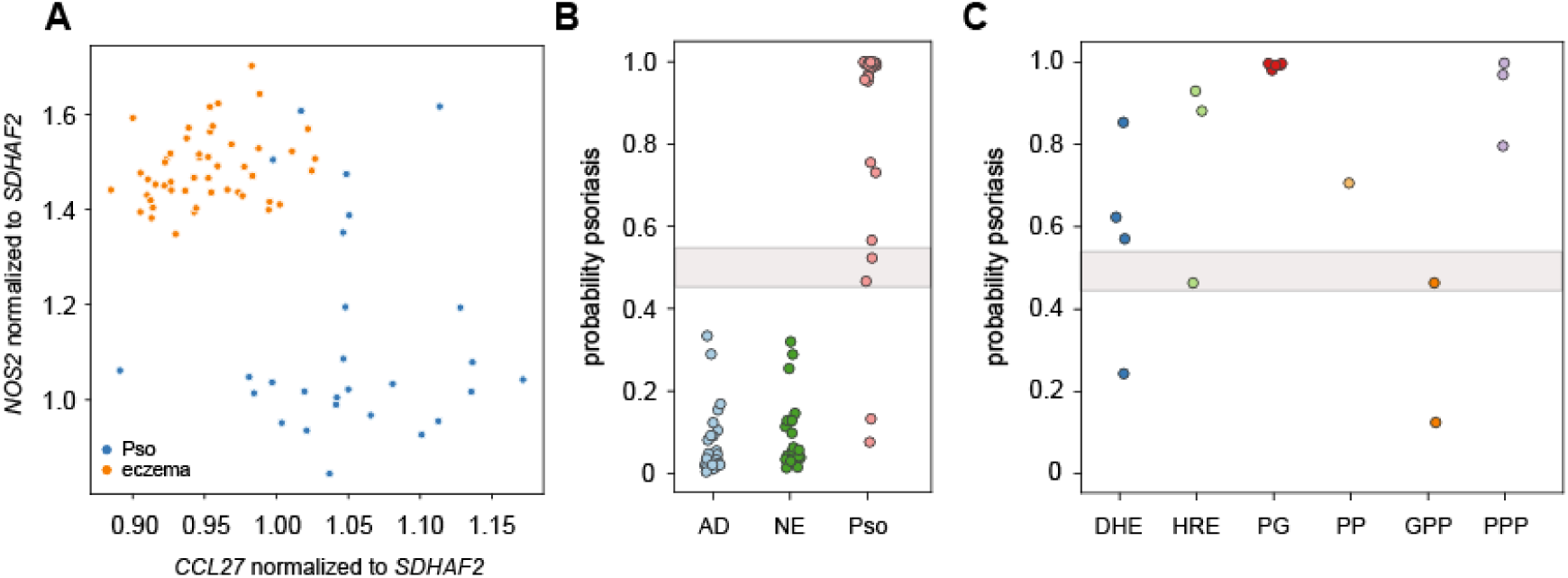
The FFPE-based Molecular classifier (MC) dissects clinically clear cases of psoriasis and eczema and is also efficient in subtypes of both diseases. A) The molecular classifier (MC) was trained on clear cases of eczema (atopic dermatitis (AD n=28), nummular eczema (NE, n=21) and psoriasis (Pso, n=27) using a logistic regression model with a 5-fold cross validation delivering the probability of psoriasis. CT values of *NOS2* and *CCL27* were normalized to the reference genes *SDHAF2* and *TBP*. The graph indicates the dissection of eczema and psoriasis samples exemplarily for normalization B) FFPE-based MC determined probabilities for psoriasis in clear cases of atopic dermatitis (AD), nummular eczema (NE) and psoriasis (Pso). The grey are between 45%-55% indicates undetermined probabilities C) The FFPE-based MC was used to predict the probability for psoriasis in subtypes of eczema and psoriasis (dyshidrotic eczema, DHE n=; hyperkeratotic-rhagadiform eczema, HRE n=; psoriasis guttata, PG n=5; psoriasis palmoplantaris, PP n=1); generalized pustular psoriasis, GPP n=2; and psoriasis pustulosa palmplantaris, PPP n=2). A probability between 45% and 55% was defined as not evaluable.

### *FFPE classifier results correlate with clinical attributes* of psoriasis and eczema

To understand if the determined probability for psoriasis also correlates with clinical attributes associated with psoriasis or eczema, Person correlation coefficients were determined (Fig. 2). The probability of psoriasis highly and positively correlated with the histological attributes ’dilated dermal capillaries’ (r=0.61), ‘acanthosis’ (r=0.51), ‘neutrophils’ (r=0.39) and ‘parakeratosis’ (r=0.33) (Fig. 2) – all hallmarks for the pathogenesis of psoriasis. This positive correlation was followed by a positive correlation of negative CT values of *NOS2* (r=.43; r=0.33; r=0.23; r=0.25, respectively) and a negative correlation with negative delta CT values for *CCL27* (r=-0.54; r=-0.5; r=-0.48; r=-0.22, respectively). In line, the probability of psoriasis and the negative delta CT for *NOS2* correlated positive with a psoriasis type morphology (r=0.77 and r=0.56) and arthritis (r=0.34 and r=0.35), whereas a negative correlation was determined for an eczema type morphology (r=-0.77 and r=-0.56) and rhinoconjunctivitis allergica (RCA) (r=-0.47 and r=-0.28) (Fig. 2). The probability of psoriasis also correlated positive with the therapeutic efficiency of IL12/IL-23 (r=0.99) and TNF-a inhibition (r=0.56), however negative with the response to Fumarate (r=-0.34) of IL-17 inhibition (r=-0.74). No correlations of the laboratory parameters ‘Stap.aureus’ and specific ‘IgE’ as well as the family history of for psoriasis and eczema were identified.

**Figure 2:**
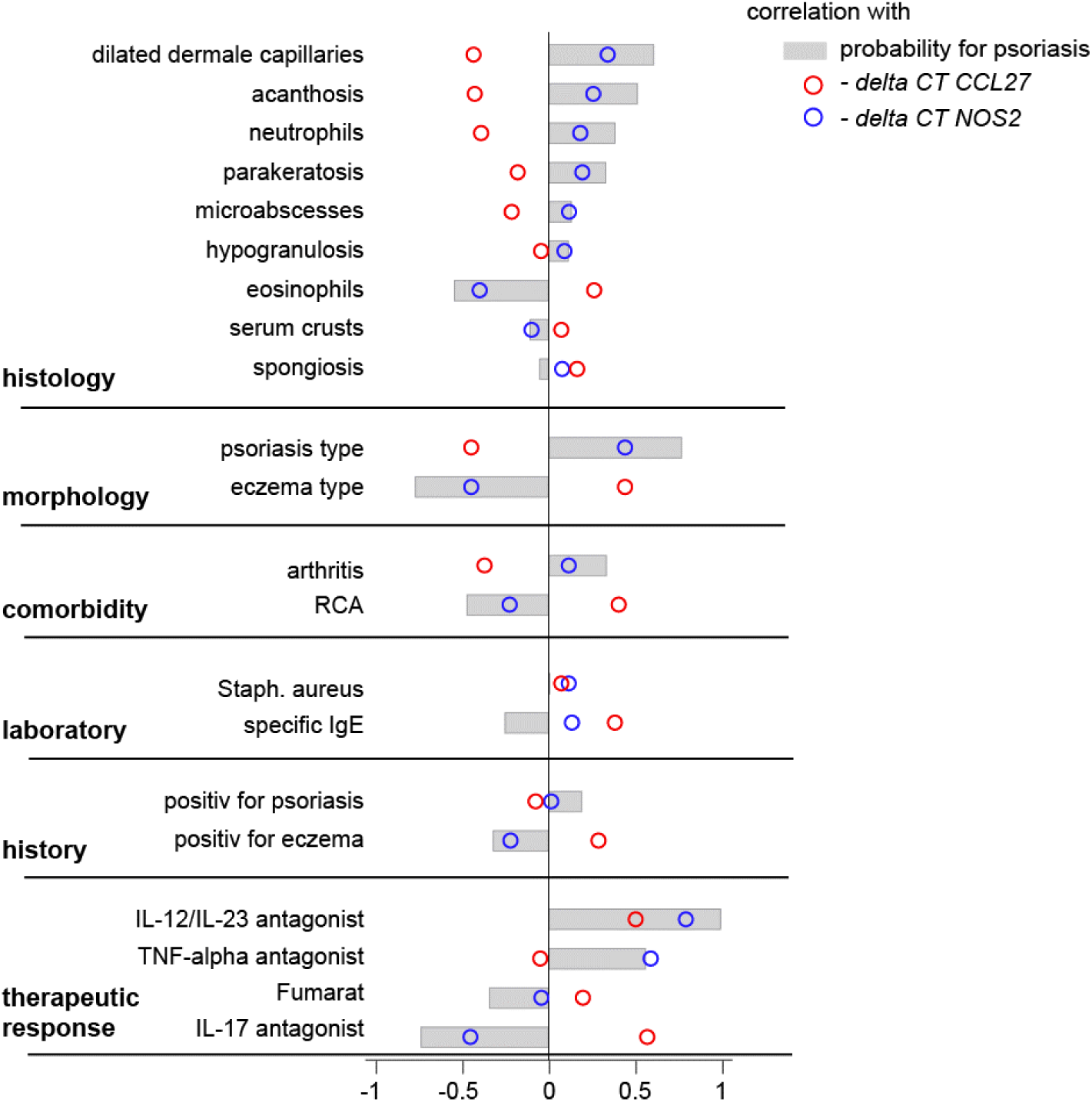
The FFPE-based MC determined probability for psoriasis correlates with disease hallmarks. The probability for psoriasis as well as the negative delta CT values for CCL27 and NOS2 were correlated with clinical attributes by Pearson correlation.

### The FFPE-based MC supports diagnostic decision making in patients with inconsistent diagnosis

To further evaluate the potential of the FFPE-based MC to support diagnostic decision making, 14 patient samples with discrepant clinical and histological diagnosis were analyzed. Within this cohort, 9 patients were assigned a probability for psoriasis and 5 patients were classified as eczema (Fig. 3A). Patient 43, a close to 80 year old woman, developed shortly after a respiratory infection itchy, pustular reddening overall the whole body. Bacterial or viral infections were not detected. Extensive lesional skin at the thigh and knee were surgically excised. Histologically, the lesions presented with infiltration of neutrophils and isolated eosinophils, signs of spongiosis and in part parakeratosis. The patient was diagnosed with psoriasis pustulosa, however, acute generalized exanthematic pustulosis (AGEP) could not be ruled out. In fact, psoriasis pustulosa and AGEP can hardly be distinguished by classical histological examination. Our FFPE-based MC assigned this patient a probability of psoriasis of 74.49%. After initiation of Ixekizumab, the skin status of this patient improved supporting the MC-based classification (Fig. 3B).

**Figure 3:**
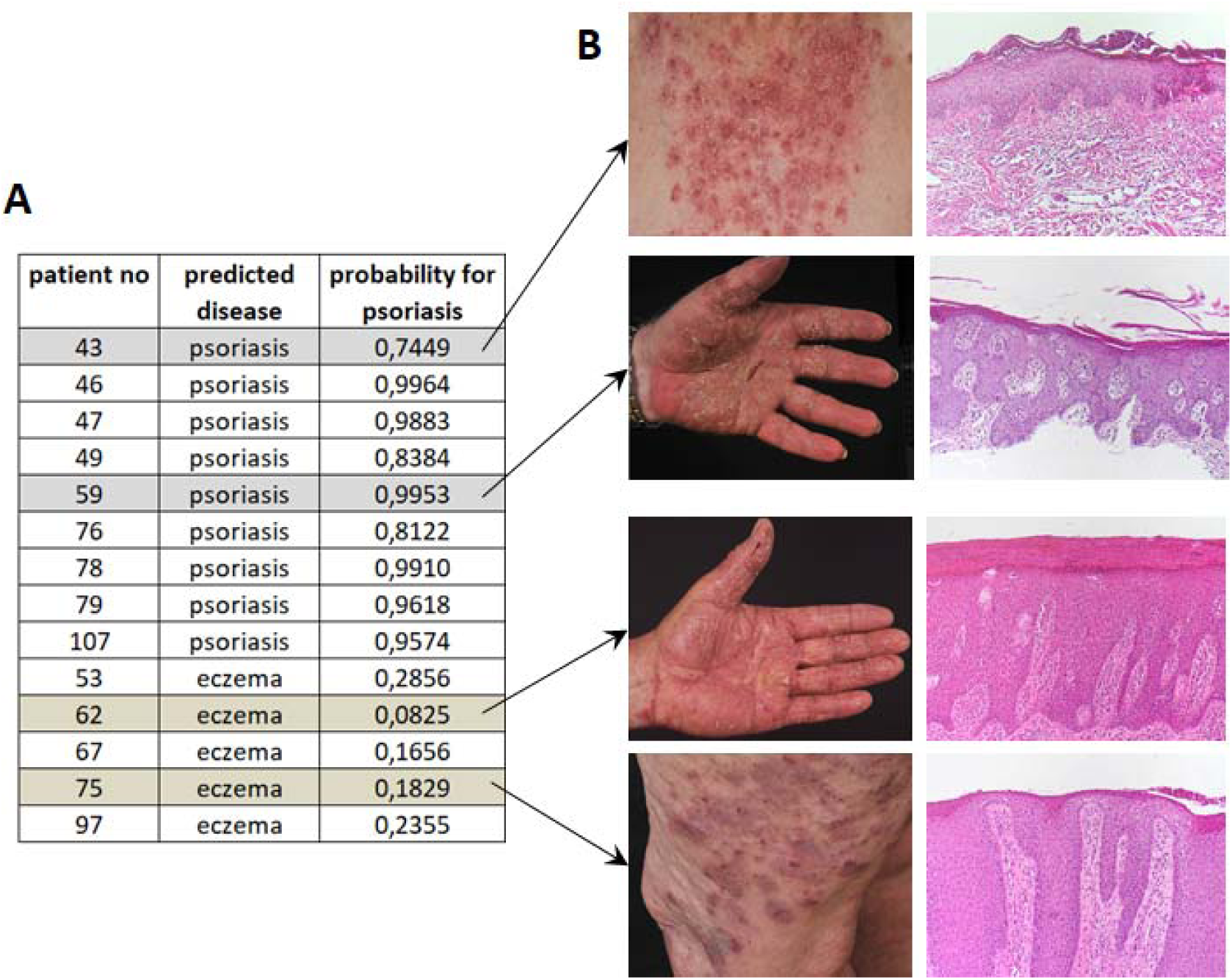
The FFPE-based MC is able to support diagnosis of unclear cases. (A) The FFPE-based MC was applied on clinically and histologically challenging cases of patients with a suspected diagnosis of psoriasis or eczema and subtypes of both diseases (n=14). Given patient numbers represent numerical numbering and not biobank IDs. B) Clinical pictures and histological H&E stainings of selected patients that were assigned a probability of psoriasis by the FFPE-based MC.

Patient 59, a mid-sixty male, presented in the clinic with reddish, hyperkeratotic, scaly and itchy skin lesions at both palmae accompanied by erythematous, heavily infiltrated plaques at the back, the capillitium and retroauricular. Histopathological examination showed psoriasiform acanthosis, hyperparakeratosis, a lymphocytic infiltrate and isolated plasma cells leading to the diagnosis of psoriasiform perivascular dermatitis, while clinically diagnostics was in favor for psoriasis. The FFPE-based MC assigned a probability for psoriasis of 99.53%. Skin lesions at the palms and body cleared after 6 months of methothrexate therapy (Fig. 3B).

Patient 62, a male at the end of his sixties, presented at the clinic with confluent, erythematous, crude-lamellar, scaly plaques at both palmae. Histologically, extended hyperparakeratosis, serum crusts, psoriasiforme epidermis hyperplasia combined with spongiosis and a linear lymphocytic infiltrate in absence of neutrophils and eosinophils was described leading to the diagnosis of hyperkeratotic, in part dyshidrotic hand eczema. The FFPE-based MC assigned this patient a probability for psoriasis of 8.25% and thereby a high likelihood of an underlying eczematous disease. Treatment with a topical corticosteroids and altitretinoin improved the skin lesions supporting the MC-based classification (Fig. 3B).

Patient 75, a woman at beginning of her sixties, has been suffering for years under recurrent itchy, sharply demarcated erythematous plaques, with in part lamellar scales and excoriations as clinically seen in both psoriasis and eczema variants. Histologically, a psoriasiform hyperplasia of the epidermis combined with hyperparakeratosis, hypogranulosis, elongated rete-ridges, papillomatosis and focal spongiosis in upper epidermis layers was found. A perivascular dense lymphocytic infiltrate with absence of neutrophils and eosinophils was identified. The patient was diagnosed with a nummular eczema. This is supported also by the FFPE-based MC that assigned a probability of 18.29% to this patient (Fig. 3B) and skin lesions improved with topical corticosteroids and UVB phototherapy.

## Discussion

The development of highly specific biologic drugs has revolutionized treatment options for various diseases during the past two decades (Noda, Krueger, & Guttman-Yassky, 2015). However, due to their specificity, these drugs are most efficient in only a small group of diseases and patients calling for efficient diagnosis and subsequent assignment of each patient to the most appropriate drug. Whereas molecular diagnostics have been already developed and used for cancer diagnosis followed by decision making on treatment regimens, they are not well established in the field of chronic inflammatory skin diseases, yet (Grazia, Penna, Perotti, Anichini, & Tassi, 2014; Levine & Fisher, 2014; Usher-Smith, Emery, Kassianos, & Walter, 2014). In previous work, we have developed a disease classifier to dissect psoriasis from eczema based on the expression levels of *NOS2* and *CCL27* (Quaranta et al., 2014; Garzorz-Stark et al., 2016). This classifier has been validated by several groups and found its acceptance in the field (Bentz et al., 2021; Brunner et al., 2018; Stoffel et al., 2018). However, it was established on RNA-later fixed skin that is regularly used in research, but not in clinics, private practices and pathology laboratories. Within these institutions, skin samples usually are formalin fixed and paraffin embedded (FFPE). This way of fixation allows long-term storage of samples and optimal assessment of histological hallmarks, however, by cross-linking of proteins with RNA and RNA degradation due to the fixation procedure, RNA isolation from FFPE samples can be challenging. Here, optimized protocols have been developed to minimize side effects of the fixation method and now allow to use this sample type for efficient diagnostics. Within this work, we have transferred the RNA-later molecular classifier to FFPE samples and were able to retrieve comparable results. Whereas the RNA-later based MC dissected psoriasis from eczema with a specificity and sensitivity for psoriasis of 100% and 97.7%, respectively and an AUC of 0.99, the FFPE-based classifier determined probabilities for psoriasis with a specificity and sensitivity of 100% and 92%, respectively, and an AUC of 0.97 (Garzorz-Stark et al., 2016). Whereas only two classical psoriasis samples were not classified correctly by the FFPE-based MC, all atopic dermatitis and nummular eczema cases were diagnosed precisely. The FFPE-based MC was also able to predict the diagnosis of psoriasis subtypes, however, delivered conflicting results for dyshidrotic and hyperkeratotic-rhagadiform eczema. Unfortunately, the cohort of these subtypes was very small (4 and 3 cases, respectively) and data on treatment regimens and therapeutic efficiency were not available. As these patients are difficult to diagnose by gold standard diagnostics, further studies need to be undertaken to prove the efficiency of molecular classification in this eczema subtype. In addition, and in line with previous findings, the FFPE-based classifier probability highly correlated with disease hallmarks of psoriasis, but not hallmarks of eczema (Garzorz-Stark et al., 2016). Therefore, the reliability of the two markers, *NOS2* and *CCL27*, has been shown now also in a FFPE tissue arguing for implementation of this molecular diagnostic aid in clinical routine diagnostics and pathology laboratories. Here, however, further efforts have to be undertaken to simplify and speed up the labor- and cost-intensive purification of RNA from FFPE tissue. In addition, fully automated systems need to be developed to be integrated into the processes in routine diagnostic laboratories.

## Data Availability

Generated data and codes can be provided upon request

## Acknowledgments

The authors thank Kerstin Weber for excellent technical support and the Biobank Biederstein for patient samples. This work was supported by the Medical Valley Award.

## References

Bentz, P., Eyerich, K., Weber, K., Kluge, L., Ofenloch, R., & Weisshaar, E. (2021). [Cohort study for long-term follow-up of patients in whom the so-called “molecular classifier” is used to distinguish eczema from psoriasis: Background and implementation]. Hautarzt, 72(4), 354–357. doi:10.1007/s00105-021-04774-9

Brunner, P. M., Pavel, A. B., Khattri, S., Leonard, A., Malik, K., Rose, S., … Guttman-Yassky, E. (2018). Baseline IL22 expression in atopic dermatitis patients stratifies tissue responses to fezakinumab. J Allergy Clin Immunol. doi:10.1016/j.jaci.2018.07.028

Bylund, S., von Kobyletzki, L. B., Svalstedt, M., & Svensson, A. (2020). Prevalence and Incidence of Atopic Dermatitis: A Systematic Review. Acta Derm Venereol, 100(12), adv00160. doi:10.2340/00015555-3510

Garzorz-Stark, N., Krause, L., Lauffer, F., Atenhan, A., Thomas, J., Stark, S. P., … Eyerich, K. (2016). A novel molecular disease classifier for psoriasis and eczema. Exp Dermatol, 25(10), 767–774. doi:10.1111/exd.13077

Grazia, G., Penna, I., Perotti, V., Anichini, A., & Tassi, E. (2014). Towards combinatorial targeted therapy in melanoma: from pre-clinical evidence to clinical application (review). Int J Oncol, 45(3), 929–949. doi:10.3892/ijo.2014.2491

Kolesnik, M., Franke, I., Lux, A., Quist, S. R., & Gollnick, H. P. (2018). Eczema in Psoriatico: An Important Differential Diagnosis Between Chronic Allergic Contact Dermatitis and Psoriasis in Palmoplantar Localization. Acta Derm Venereol, 98(1), 50–58. doi:10.2340/00015555-2779

Levine, D., & Fisher, D. E. (2014). Current status of diagnostic and prognostic markers in melanoma. Methods Mol Biol, 1102, 177–197. doi:10.1007/978-1-62703-727-3_11

Noda, S., Krueger, J. G., & Guttman-Yassky, E. (2015). The translational revolution and use of biologics in patients with inflammatory skin diseases. J Allergy Clin Immunol, 135(2), 324–336. doi:10.1016/j.jaci.2014.11.015

Quaranta, M., Knapp, B., Garzorz, N., Mattii, M., Pullabhatla, V., Pennino, D., … Eyerich, K. (2014). Intraindividual genome expression analysis reveals a specific molecular signature of psoriasis and eczema. Sci Transl Med, 6(244), 244ra290. doi:10.1126/scitranslmed.3008946

Renert-Yuval, Y., Thyssen, J. P., Bissonnette, R., Bieber, T., Kabashima, K., Hijnen, D., & Guttman-Yassky, E. (2021). Biomarkers in atopic dermatitis-a review on behalf of the International Eczema Council. J Allergy Clin Immunol, 147(4), 1174–1190 e1171. doi:10.1016/j.jaci.2021.01.013

Stoffel, E., Maier, H., Riedl, E., Bruggen, M. C., Reininger, B., Schaschinger, M., … Brunner, P. M. (2018). Analysis of anti-tumour necrosis factor-induced skin lesions reveals strong T helper 1 activation with some distinct immunological characteristics. Br J Dermatol, 178(5), 1151–1162. doi:10.1111/bjd.16126

Thyssen, J. P., Johansen, J. D., Linneberg, A., & Menne, T. (2010). The epidemiology of hand eczema in the general population--prevalence and main findings. Contact Dermatitis, 62(2), 75–87. doi:10.1111/j.1600-0536.2009.01669.x

Tizek, L., Schuster, B., Gebhardt, C., Reich, K., von Kiedrowski, R., Biedermann, T., … Garzorz-Stark, N. (2021). Molecular diagnostics in dermatology: An online survey to study usage, obstacles and requirements in Germany. J Dtsch Dermatol Ges. doi:10.1111/ddg.14659

Usher-Smith, J. A., Emery, J., Kassianos, A. P., & Walter, F. M. (2014). Risk prediction models for melanoma: a systematic review. Cancer Epidemiol Biomarkers Prev, 23(8), 1450–1463. doi:10.1158/1055-9965.EPI-14-0295

Vena, G. A., Altomare, G., Ayala, F., Berardesca, E., Calzavara-Pinton, P., Chimenti, S., … Cricelli, C. (2010). Incidence of psoriasis and association with comorbidities in Italy: a 5-year observational study from a national primary care database. Eur J Dermatol, 20(5), 593–598. doi:10.1684/ejd.2010.1017

